# First SARS-CoV-2 detection in river water: implications in low sanitation countries

**DOI:** 10.1101/2020.06.14.20131201

**Authors:** Laura Guerrero-Latorre, Isabel Ballesteros, Irina M. Villacrés, M. Genoveva Granda, Byron P. Freire, Blanca Ríos-Touma

**Affiliations:** Grupo de Investigación en Biodiversidad, Medio Ambiente y Salud. Facultad de Ingenierías y Ciencias Aplicadas. Universidad de Las Américas, Ecuador; Department of Biology, Area of Microbiology, University of Girona, Spain; Laboratorios de Investigación. Universidad de Las Américas, Ecuador

**Keywords:** COVID-19, Andean Rivers, wastewater, viral dissemination, Ecuador, HadV

## Abstract

Since the beginning of SARS-CoV-2 pandemic studies on viral shedding have reported that this virus is excreted in feces in most patients. High viral loads are found at the sewage pipeline or at the entrance of wastewater treatment plants from cities where the number of COVID-19 cases are significant. In Quito (Ecuador) as in many other cities worldwide, wastewater is directly discharged into natural waters. The aim of this study was to evaluate SARS-CoV-2 presence in urban streams from a low sanitation context. Three river locations along the urban rivers of Quito were sampled on the 5th of June during a peak of COVID-19 cases. River samples were evaluated for water quality parameters and afterwards, concentrated for viral analysis using skimmed milk flocculation method. The viral concentrates were quantified for SARS-CoV-2 (N1 and N2 genes) and Human Adenovirus as a human viral indicator. The results showed that SARS-CoV-2 was detected for both target genes in all samples analyzed in a range of 2,91E+05 to 3,19E+06 GC/L for N1 and from 2,07E+05 to 2,22E+06 GC/L for N2. The high values detected in natural waters from a low sanitation region have several implications in health and ecology that should be further assessed.

## 1. Introduction

Since the first detected case of unknown pneumonia in Wuhan (China), in half a year, SARS-CoV-2 has caused almost 8 million confirmed cases and more than 400.000 deaths worldwide (WHO, 2020). Currently, Latin America is considered the epicenter of the pandemic (UN, 2020). Brazil reported Latin America’s first case in late February 2020, and since then, COVID-19 cases and deaths have increased widely throughout nearly every country in the region. In Ecuador, official reports declare more than 45.000 cases and 3.800 deaths in the second week of june (SNGRE, 2020).

SARS-CoV-2 transmission in humans is mainly airborne (Zhang *et al*., 2020) but recent studies suggest that extended shedding of SARS-CoV-2 in feces is a potential health risk (Lodder and Roda Husman, 2020). As most COVID-19 cases fecally excreted the virus, SARS-CoV-2 has been detected in wastewater treatment plants from several cities around the world and some countries have started to monitor the viral load in sewage systems in an attempt to design early warnings for future outbreaks (Bivins *et al*., 2020).

However, the fecal shedding in wastewater could be especially relevant in low sanitation countries where insufficient or nonexistent treatments of wastewater are applied.

Studies analyzing SARS-CoV-2 in sewage have tested the efficiency after wastewater treatments applied and in most cases effluent waters did not show the presence of SARS-CoV-2 (Randazzo *et al*., 2020a). Moreover, in Japan, Haramoto et al. 2020 looked for SARS-CoV-2 in rivers impacted by sewage, and did not detect its presence. In most countries of Latin America an average of 30% of wastewaters are treated before discharging in water bodies (Rodriguez et al., 2020). In particular, Ecuador has less than 20% of treatment coverage for its wastewaters (Rodriguez et al., 2020) and its capital city, Quito with almost 3 million inhabitants only treats 3% of its sewage (EPMAPS, 2020). Therefore, the urban rivers of Quito, whose receive the fecal waters from the whole city, are highly contaminated by human microorganisms and other pollutants, disseminating them along the river basin and crossing from up 2800 masl to the Pacific coast (Guerrero-Latorre et al., 2018; Voloshenko et al 2015). Previous studies have described the viral diversity of human pathogens present in the urban rivers of Quito and highlighting the health risk of populations that can use downstream waters for irrigation or recreational purposes (Guerrero-Latorre et al 2018). In the context of the COVID-19 pandemic, very little information has emerged from low sanitation countries about the presence of SARS-Cov-2 in sewage and natural waters. Therefore, this is the first study in a low sanitation country that attempted to analyze the presence of SARS-CoV-2 in river waters impacted by urban wastewater.

## 2. Material and methods

### 2.1 Sampling

The 5th of June 2020 three locations from Quito’s river were sampled. Sampling locations are representative from south-center (M1), north-center (M2) and north (M3) accumulated wastewater discharges into the natural streams passing through the city (Figure 1, EPMAPS, 2011). At each location, 2 L samples were collected for viral analysis and 2 L for other biological and chemical measurements:DBO_5_, DQO, pH and conductivity. Samples were preserved at 4 °C and taken to the laboratory in less than 3 hours.

**Figure 1.**
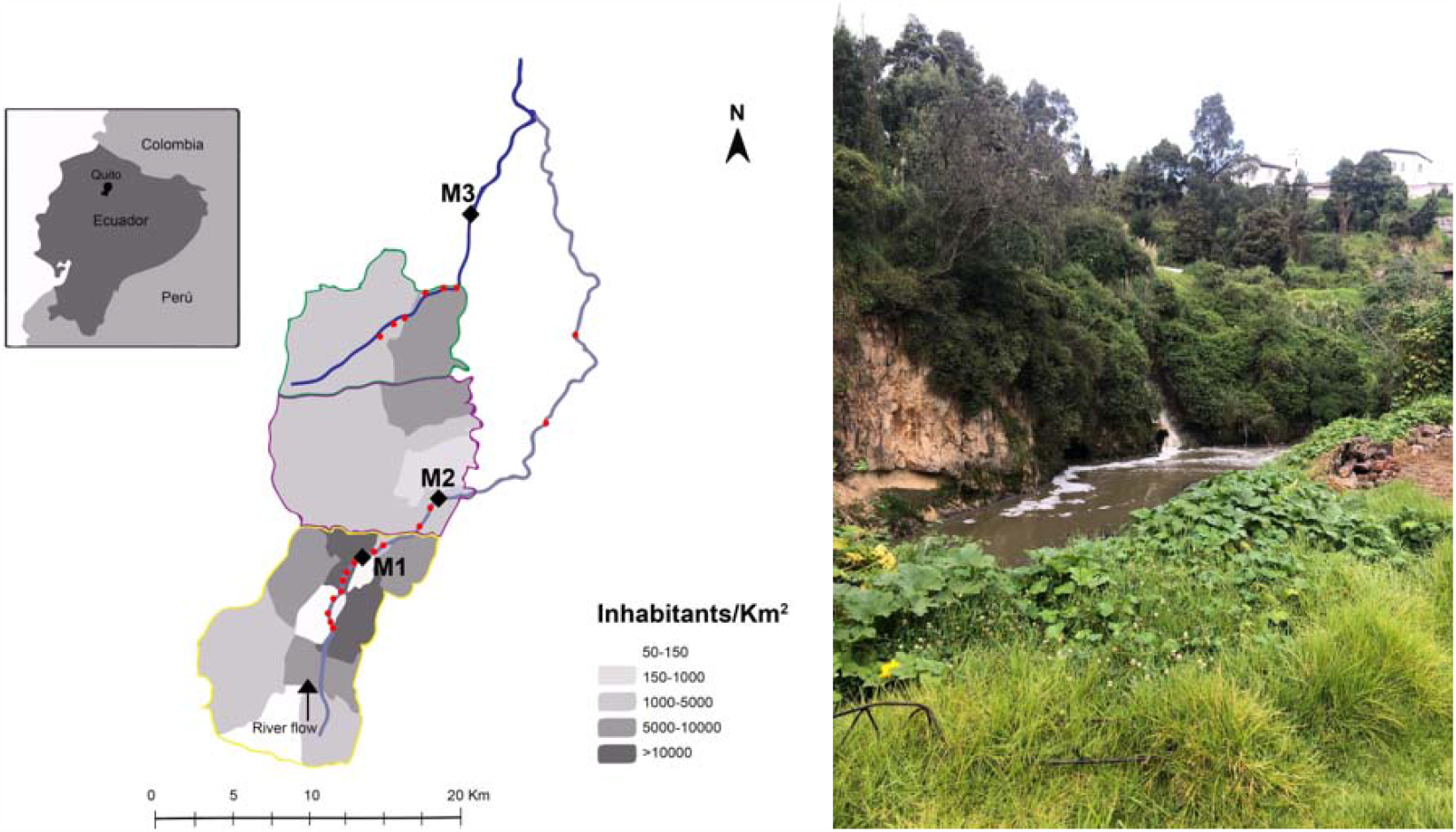
**Left**. Sampling locations in urban rivers from Quito (Guerrero-Latorre *et al*. 2018). M1 (0°,14’00.27’’S; 78°30’42.43’’O), M2 (0°11’52.47’’S;78°28’06.46’’O), M3 (0°00’42.98’’S; 78°26’25.86’’O). **Right**. Picture of direct discharge of wastewater into Machángara River (sampling point M1).

### 2.2 Water concentration and RNA extraction

Collected water was concentrated for viral analysis using an adapted Skimmed Milk Flocculation method using 2 L of water (Fernandez-Cassi *et al*., 2018). Briefly, sample was preconditioned at pH 3.5 and a conductivity above 1.5 mS/cm2. Then, 10 ml of pre-flocculated skimmed milk solution were added. After 8 h of stirring, flocks were centrifuged at 8,000× g for 40 min, and the pellet was suspended in 10 mL of phosphate buffer. Nucleic acids were extracted using AccuPrep® Universal RNA Extraction Kit - Bioneer from 200 ul of the viral concentrates into 40 ul of eluted solution.

### 2.3 qRT-PCR analysis

Nucleic acid extractions from viral concentrates were used to analyze by RT-qPCR SARS-CoV-2 using N1 and N2 primer sets (CDC, 2020) and Human Adenovirus as a microbial indicator of human fecal contamination (Allard *et al*., 2001).

The volume of original water sample analyzed in each PCR reaction after concentration and extraction steps is 3 µL for SARS-CoV-2 using TaqMan Fast Virus 1-Step Master Mix (ThermoFisher) and 10 µL for HAdV using Environmental TaqMan MasterMix (ThermoFisher). Quantitative PCRs were run in Bio-Rad CFX96 qPCR thermocycler. The inhibition was analysed by testing the direct and 1/10 diluted extractions for each qPCR or RT-qPCR.

### 2.4 Cases per sampling point

In order to relate SARS-CoV-2 viral load in river water samples with reported COVID-19 cases in Quito, we gathered information from the sectors of Quito that discharge sewage to the sampling points according to the Potable Water and Sanitation Enterprise of Quito (EPMAPS, 2011). Since the city of Quito reports its cases by these sectors we gather current and historical information from 14 days prior to the sampling points (https://twitter.com/MunicipioQuito/). This information was validated with the official government reports (SNGRE, 2020).

## 3. Results

Water quality parameters of samples analyzed show an important anthropogenic impact in the rivers (Table 1). Sampling points M2 and M3 do not meet the national requirement for aquatic life preservation for DQO and DBO_5_ (Ministerio del Ambiente de Ecuador, 2015). Human Adenovirus, a viral indicator used to evaluate the microbial impact in river water present high quantities corresponding to strong impact (Rusiñol *et al*. 2014)

**Table 1.** Water quality parameters at sampling locations

| Sampling points | pH | Conductivity | DBO<sub>5</sub> mg O<sub>2</sub>/L | DQO mg O<sub>2</sub>/L | HAdV GC/L |
| --- | --- | --- | --- | --- | --- |
| M1 | 7,73 | 653,07 | 13 | 35.706 | 1,13E+04 |
| M2 | 8,16 | 703,2 | 28* | 63.353* | 2,18E+04 |
| M3 | 7,83 | 696,48 | 16 | 43.941* | 2,60E+05 |

\*Parameters that do not comply with the Ecuadorian normative for aquatic life
preservation (Ministerio del Ambiente de Ecuador, 2015).

SARS-CoV-2 genes (N1 and N2) were present in the three locations and show concentrations ranging from 2,84E+05 to 3,19E+06 GC/L for N1 target gene and from 2,07E+05 to 2,23E+06 for N2 target gene (Table 2).

**Table 2.** Viral loads of SARS-CoV-2 per location and cumulative and active COVID-19 cases in the influence area.

| **Sampling**<br>**points** | **SARS-CoV-2 N1**<br>**GC/L** | **SARS-CoV-2 N2**<br>**GC/L** | **Cumulative cases**<br>**(total COVID-19**<br>**cases until 5th**<br>**June)** | **Active cases**<br>**(COVID-19**<br>**cases from the**<br>**prior 14 days)** |
| --- | --- | --- | --- | --- |
| M1 | 3,19E+06 | 2,23E+06 | 2077 | 579 |
| M2 | 2,84E+05 | 2,07E+05 | 580 | 81 |
| M3 | 2,91E+06 | 8,55E+05 | 358 | 90 |

These values are clearly related to COVID-19 cases reported in the contributing areas for each point, showing higher values where higher active cases were registered (Table 2). The collection of samples was during a peak of the outbreak, since the active cases (considered the notified cases in the last 14 days prior to the sampling day) represent 25% of the total cases of COVID19 reported in the city since the beginning of the outbreak.

## 4. Discussion

This is the first study that reports SARS-CoV2, and in important quantities, in rivers, however other studies have attempted to find it without success (Haramoto *et al*., 2020). The urban river of Quito is impacted by the direct discharge of sewage water from a population of almost 3 million inhabitants. The water quality parameters described in Table 1 indicate the high levels of organic and inorganic contamination from those urban rivers. The water parameters measured in 2 of the 3 locations studied were above the national regulations for aquatic life preservation (Ministerio del Ambiente de Ecuador, 2015). However, all environmental values are higher than those registered for reference sites in this same basin (Ríos-Touma *et al*., 2014)

Viral human contamination analyzed by HAdV indicator shows the great impact of human excreta in the urban rivers of Quito. Values are very similar from the samples collected in the same locations in 2017 (Guerrero-Latorre *et al*., 2018). In that study, a metagenomic analysis was carried out in the same locations and sequences from 26 human viral pathogens were obtained, revealing the microbial consequences of sewage discharge without previous treatment into natural streams.

Nowadays, in the peak of COVID-19 pandemic in Ecuador, levels of SARS-CoV-2 found in urban rivers of Quito are similar as found in sewage of Valencia (Spain) with more than 5000 active cases (Randazzo *et al*. 2020b) and Paris in the epidemic peak of cases with more than 10000 hospitalized cases (Wurtzel *et al*., 2020). However, at the sampling date in the urban area of Quito only 750 COVID-19 active cases were notified, suggesting an important underdiagnose as seen in many other regions worldwide.

The implications from our study can be extended to other cities where the sewage is directly discharged into natural streams. Firstly, SARS-CoV-2 presence as other microbial pathogens discharged in open river water might pose a risk of infection for the population in contact with the water downstream. The risk has been recently approached in a QMRA study of WWTP workers exposed to a different concentration of SARS-CoV-2 in sewage (Zaneti *et al*., 2020). However, it is important to remark that only genomic material has been detected in waters and the viability of the virus is not known for polluted waters. Secondly, the viral dissemination into the environment has an unknown impact into livestock and wildlife health, as the spillover events of zoonotic links are frequent in the coronaviridae family (Franklin and Bevins, 2020).

Finally, the presence of the virus can be also used as a surveillance tool for an early warning system using main sewage discharges along the city helping to control the pandemic where diagnostic tools are limited (Bivins *et al*., 2020). Also, the method implemented in the present publication can be used in other cities where sewage is not possible to sample and wastewaters are discharged to streams or rivers.

## 5. Conclusions

We have detected for the first time important viral loads of SARS-CoV-2 from rivers in urban streams of Quito. The loads found suggest that cases are probably a lot higher than the official data. Also, implications in dissemination of SARS-CoV-2 in low sanitation countries by polluted freshwaters should be further considered. The low degree of wastewater treatment coverage in the region, might be a factor of increased risk for COVID-19 pandemic.

## Data Availability

Data is included in the manuscript

## Acknowledgements

Special thanks to Xavier Amigo and Nature Experience for its invaluable service during the sampling campaign.

## Fuding

This work was supported by research funds from Universidad de Las Américas, Ecuador.

## References

Allard, A., Albinsson, B., Wadell, G., Albinsson, B.O., 2001. Rapid Typing of Human Adenoviruses by a General PCR Combined with Restriction Endonuclease Analysis Rapid Typing of Human Adenoviruses by a General PCR Combined with Restriction Endonuclease Analysis. J. Clin. Microbiol. 39, 498–505. https://doi.org/10.1128/JCM.39.2.498

Bivins, A., North, D., Ahmad, A., Ahmed, W., Alm, E., Been, F., et al, 2020. Wastewater-Based Epidemiology: Global Collaborative to Maximize Contributions in the Fight Against COVID-19. Environ. Sci. Technol. doi:10.1021/acs.est.0c02388

CDC, 2020. 2019-Novel Coronavirus (2019-nCoV) Real-Time RT-PCR Diagnostic Panel. US Centers for Disease Control and Prevention. https://www.cdc.gov/coronavirus/2019-ncov/lab/rt-pcr-panel-primer-probes.html [Accessed 3 Mar 2020].

Empresa Pública Metropolitana de Agua Potable y Saneamiento. (2011). Estudios de Actualización del Plan Maestro Integrado de Agua Potable y Alcantarillado para el DMQ. Recuperado el 4 de febrero del 2018 de https://www.aguaquito.gob.ec/sites/default/files/documentos/plan_maestro_agua_potable.pdf

Empresa Pública Metropolitana de Agua Potable y Saneamiento. (2020). Programa para la descontaminación de los ríos de Quito. https://www.aguaquito.gob.ec/programa-para-la-descontaminacion-de-los-rios-de-quito/. [Accessed June 13th, 2020]

Fernandez-Cassi X., Timoneda, N., Martínez-Puchol, S., Rusiñol, M., Rodriguez-Manzano, J., Figuerola, N., et al 2018. Metagenomics for the Study of Viruses in Urban Sewage as a Tool for Public Health Surveillance. Sci Total Environ. Mar 15;618:870–880. https://doi.org/10.1016/j.scitotenv.2017.08.249

Franklin, A. B., & Bevins, S. N. (2020). Spillover of SARS-CoV-2 into novel wild hosts in North America: A conceptual model for perpetuation of the pathogen. The Science of the total environment, 733, 139358. Advance online publication. https://doi.org/10.1016/j.scitotenv.2020.139358

Guerrero-Latorre, L., Romero, B., Bonifaz E., Timoneda, N., Rusiñol, M., Girones, R. and Rios-Touma, B. 2018. Quito’s virome: Metagenomic analysis of viral diversity in urban streams of Ecuador’s capital city. Sci Total Environ. 645, pp. 1334 – 1343; doi:https://doi.org/10.1016/j.scitotenv.2018.07.213

Haramoto, E., Malla, B., Thakali, O., Kitajima, M., 2020. First environmental surveillance for the presence of SARS-CoV-2 RNA in wastewater and river water in Japan. medRxiv 2020.06.04.20122747; doi: https://doi.org/10.1101/2020.06.04.20122747

Lodder, W., de Roda Husman, A.M., 2020. SARS-CoV-2 in wastewater: potential health risk, but also data source. Lancet Gastroenterol. Hepatol. 5, 533–534. https://doi.org/10.1016/S2468-1253(20)30087-X

Ministerio del Ambiente de Ecuador. 2015. 097-A Refórmese el Texto Unificado de Legislación Secundaria. Registro Oficial. Año III - No 387.Quito, miércoles 4 de noviembre de 2015. P 6 a 78.

Randazzo, W., Truchado, P., Cuevas-Ferrando, E., Simon, P., Allende, A. and Sanchez, G. 2020a. SARS-CoV-2 RNA in wastewater anticipated COVID-19 occurrence in a low prevalence area. Water Research. Volume 181, 15 August 2020, 115942. https://doi.org/10.1016/j.watres.2020.115942

Randazzo, W., Cuevas-Ferrando, E., Sanjuan, R., Domingo-Calap, P., Sanchez, G., 2020b. Metropolitan Wastewater Analysis for COVID-19 Epidemiological Surveillance. medRxiv 2020.04.23.20076679. https://doi.org/10.1101/2020.04.23.20076679

Ríos-Touma, B., Acosta, R., Prat, N., 2014. The Andean biotic index (ABI): Revised tolerance to pollution values for macroinvertebrate families and index performance evaluation. Rev. Biol. Trop. 62, 249–273.

Rodriguez Hector, Alexander Delgado, Anna Nolasco, Daniel Saltiel, Gustavo, D. J. S. 2020. From Waste to Resource. Water Papers. World Bank. https://doi.org/doi:10.1596/33436

Rusiñol, M., Fernandez-Cassi, X., Hundesa, A., Vieira, C., Kern, A., Eriksson, I. et al.. (2014). Application of human and animal viral microbial source tracking tools in fresh and marine waters from five different geographical areas. Water Research. 59, pp. 119–129. https://doi.org/doi:10.1016/j.watres.2014.04.013

SNGRE 2020. Informes de Situación e Infografias – COVID 19 – desde el 29 de Febrero del 2020. https://www.gestionderiesgos.gob.ec/informes-de-situacion-covid-19-desde-el-13-de-marzo-del-2020/

UN 2020. Central and South America now ‘intense zones’ for COVID-19 transmission. 1 June 2020 https://news.un.org/en/story/2020/06/1065252

Voloshenko-Rossin, A., Gasser, G., Cohen, K., Gun, J., Cumbal-Flores, L., Parra-Morales, W., Sarabia, F., Ojeda, F., Lev, O., 2015. Emerging pollutants in the Esmeraldas watershed in Ecuador: discharge and attenuation of emerging organic pollutants along the San Pedro–Guayllabamba–Esmeraldas rivers. Environ. Sci. Process. Impacts 17, 41–53. https://doi.org/10.1039/C4EM00394B

WHO. WHO Coronavirus Disease (COVID-19) Dashboard. 2020. https://covid19.who.int/

Wurtzer, S., Marechal, V., Mouchel, JM., Maday, Y., Teyssou, R., Richard, E., et al. 2020. Evaluation of lockdown impact on SARS-CoV-2 dynamics through viral genome quantification in Paris wastewaters. medRxiv 2020.04.12.20062679; doi: https://doi.org/10.1101/2020.04.12.20062679

Zaneti, R.N., Girardi, V., Spilki, F.R., Mena, K., Campos-Westphalen, A.P., da Costa Colares, W.R. et al. 2020. QMRA of SARS-CoV-2 for workers in wastewater treatment plants. medRxiv 2020.05.28.20116277; doi: https://doi.org/10.1101/2020.05.28.20116277

Zhang, R., Li, Y., Zhang, A.L., Wang, Y., Molina, M.J., 2020. Identifying airborne transmission as the dominant route for the spread of COVID-19. Proc. Natl. Acad. Sci. 202009637. doi:10.1073/pnas.2009637117

